# Ethnic and sexual identities: inequalities in adolescent health and wellbeing in a national population-based study

**DOI:** 10.1101/2021.07.12.21260381

**Authors:** Amal R. Khanolkar, David M. Frost, Evangeline Tabor, Victoria Redclift, Rebekah Amos, Praveetha Patalay

## Abstract

**Purpose:** This study employed an intersectional framework to examine impact of inequities related to sexual and ethnic minority identities in risk for health, wellbeing, and health-related behaviors in a nationally representative sample.

**Methods:** The study sample comprised 9,789 (51% female) adolescents aged 17 years from the UK-wide Millennium Cohort Study, with data on self-identified sexual- and ethnic-identities. Adolescents were grouped into White-Heterosexual, White sexual minority (White-SM), ethnic minority (EM)-heterosexual, and ethnic- and sexual minority (EM-SM) categories.

Mental health (e.g., self-reported psychological distress, doctor-diagnosed depression, attempted suicide, victimisation), general health (self-rated health, chronic illness, Body mass index) and a range of health-related behaviors (e.g., smoking, substance use) were assessed by questionnaires. Associations were analysed using logistic and linear regression (adjusted for sex and parental income).

**Results:** Sexual minority individuals (White:18% and ethnic minority:3%) had increased odds for mental ill-health and attempted suicide, with higher odds in White-SM than EM-SM. Compared to White-heterosexual individuals, White-SM and EM-SM had higher risk for psychological distress (Odds ratios [OR]3.47/2.24 for White-SM/EM-SM respectively), and emotional problems (OR3.17/1.65). They had higher odds for attempted suicide (OR2.78/2.02), self-harm (OR3.06/1.52), and poor sleep quality (OR1.88/1.67). In contrast, White-Heterosexual and White-SM groups had similarly high proportions reporting risky behaviors except for drug use (OR1.34) and risky sex (OR1.40) which are more common in White-SM individuals. EM-Heterosexual and EM-SM individuals had decreased odds for health-related behaviors.

**Conclusions:** Sexual minority (White and EM) individuals had substantially worse mental health compared to their heterosexual peers. Adverse health-related behaviors were more common in White sexual minority individuals. Investigation into the mechanisms leading to these differences is needed.

## Introduction

Recent decades have witnessed sociocultural shifts in better understanding sexual identities along with greater social acceptance and political rights for sexual minority (SM) groups in many countries.^1^ Nonetheless, growing evidence shows adolescent and adult SM (e.g., lesbian, gay, bisexual) have substantially poorer mental health, higher levels of stress and increased risk of self-harm, adverse health-related behaviors (e.g., substance use, physical activity, risky sex), and attempted suicide compared to their heterosexual peers.^1-4^ They are more likely to be bullied or harassed at schools and workplaces, face stigma and hide their sexual identities.^1, 5-7^

To date most studies on health in SM individuals have focused on mental health, wellbeing, and sexual health. They are mostly restricted to White individuals with the few studies on sexual *and* ethnic minority individuals originating in the US (often based on small study samples, non-probability samples, without comparator groups, some with discordant results and a substantial number focusing on one ethnic/racial group compared to the majority White group).^2, 8^ Additionally, most studies focus on adults with few studies specifically examining health and related issues in SM adolescents in general, irrespective of ethnic origin.^2^ This is despite adolescents coming out at earlier ages and increasing evidence for greater sexuality-based inequalities in adolescent health.^2, 9^

In the UK there is little knowledge on health and health-related behaviors in ethnic minority [EM] individuals who also identify as being SM (i.e., dual minority identities) and in general how combined minority statuses can impact health leading to disparities that may track into adulthood. This critical gap in knowledge is striking despite the substantial evidence of poorer health in many EM groups.^10-12^ EM populations in the UK continue to experience persistent health inequalities as a result of complex associations between historical racism, poverty, social deprivation, ethnic and health vulnerabilities^13^. One exception being better internalising mental health in millennial adolescent EM compared to White peers.^14^ EM individuals are more likely to face bullying, harassment, and racism because of their ethnic-origin and/or religion, regardless of their sexual identity.^6, 15^ Dual minority (based on ethnicity and sexuality) individuals may experience higher levels of stressors due to ethnic and cultural differences and expectations (e.g. having to conform to ethnic-specific expectations regarding gender roles, inability to disclose sexual orientation with families), and face higher levels of homophobic violence.^16, 17^ Higher levels of stress impact not only mental health but might lead to higher levels of risky and (mal)adaptive behaviors including coping mechanisms (positive and negative).^18^ Thus, EM groups could be at an even higher risk for mental health difficulties, poor wellbeing, and risky behavior due to the cumulative effects of two or more minority identities.

It is hypothesised that adverse health in individuals with ≥2 minority identities is due to the intersection between multiple ‘subordinated’ minority identities associated with different forms of and higher levels of discrimination within set social hierarchies resulting in multiple levels of inequality (the intersectionality framework theory).^19^ A key aspect of intersectionality is that it postulates that experiences at the intersection of identities are co-constituted and should be estimated jointly.^20^

Adolescence is characterised by rapid biological changes and increased psychosocial awareness. It is a critical period of increasing self-awareness and social exploration when adolescents explore self-identities with specific groups (e.g. ethnicity, religious, sexual)^21^ Chronic physical and mental health conditions, and health-related behaviors (e.g., drug and alcohol use) in adolescence can have long-term consequences as they impact access to higher education, the labour market, effective and positive participation in society, and track across adulthood increasing risk for morbidity.^22^ Lastly, victimisation, stigmatisation and discrimination related to minority identities experienced in adolescence exacerbate poor health and wellbeing in vulnerable individuals with long-term consequences for health, social, economic outcomes in later life.^23^

Using an intersectional framework, this study examined dual sexual- and ethnic-identities in relation to health in a nationally representative population of adolescents aged 17 years. We investigated differences in mental health, general health, and health-related behaviors in EM adolescents who identify as SM (dual minority) and in comparison, to their EM heterosexual and White-LGB and White heterosexual peers.

## Methods

### Study design and participants

This cross-sectional study uses data from the Millennium Cohort Study (MCS), a nationally representative birth cohort following children born Sept 2000-Jan 2002.^24^ A national sample of 19,517 children from across the UK were recruited to MCS and followed-up over seven sweeps (ages 9 months, 3, 5, 7, 11, 14 and 17 years). This study includes N=10,757 children from the age 17 sweep (from 10,625 families or 73.6% of the eligible sample at this sweep) and took place between January 2018-May 2019. MCS was oversampled to have higher proportions of ethnic minority participants and socioeconomically disadvantaged families (with appropriate sample weighting ensuring national representativeness). Attrition at the age 17 sweep was predicted by single-parent families, lower-income occupation and lower educational level, black ethnicity, and male sex.

Ethics approval for the MCS study was obtained from the National Research Ethics Service Committee London— Central (reference 13/LO/1786). All data are anonymised and available to researchers via the UK Data Service. Cohort members ≥16 years provided verbal consent to take part in the overall assessment, and each survey element.

### Sexual and ethnic identity

Participants were asked ‘*Which of the following options best describes how you currently think of yourself’* and could choose from eight options (Table 1). Based on responses, participants were categorised into 1. Completely heterosexual and 2. Sexual minority (mainly heterosexual/straight, bisexual, mainly gay/lesbian and completely gay/lesbian). Previous research suggests that mainly heterosexual individuals report health differences due to similar reasons as SM groups indicating that they can be combined for analysis.^25^

**Table 1.**
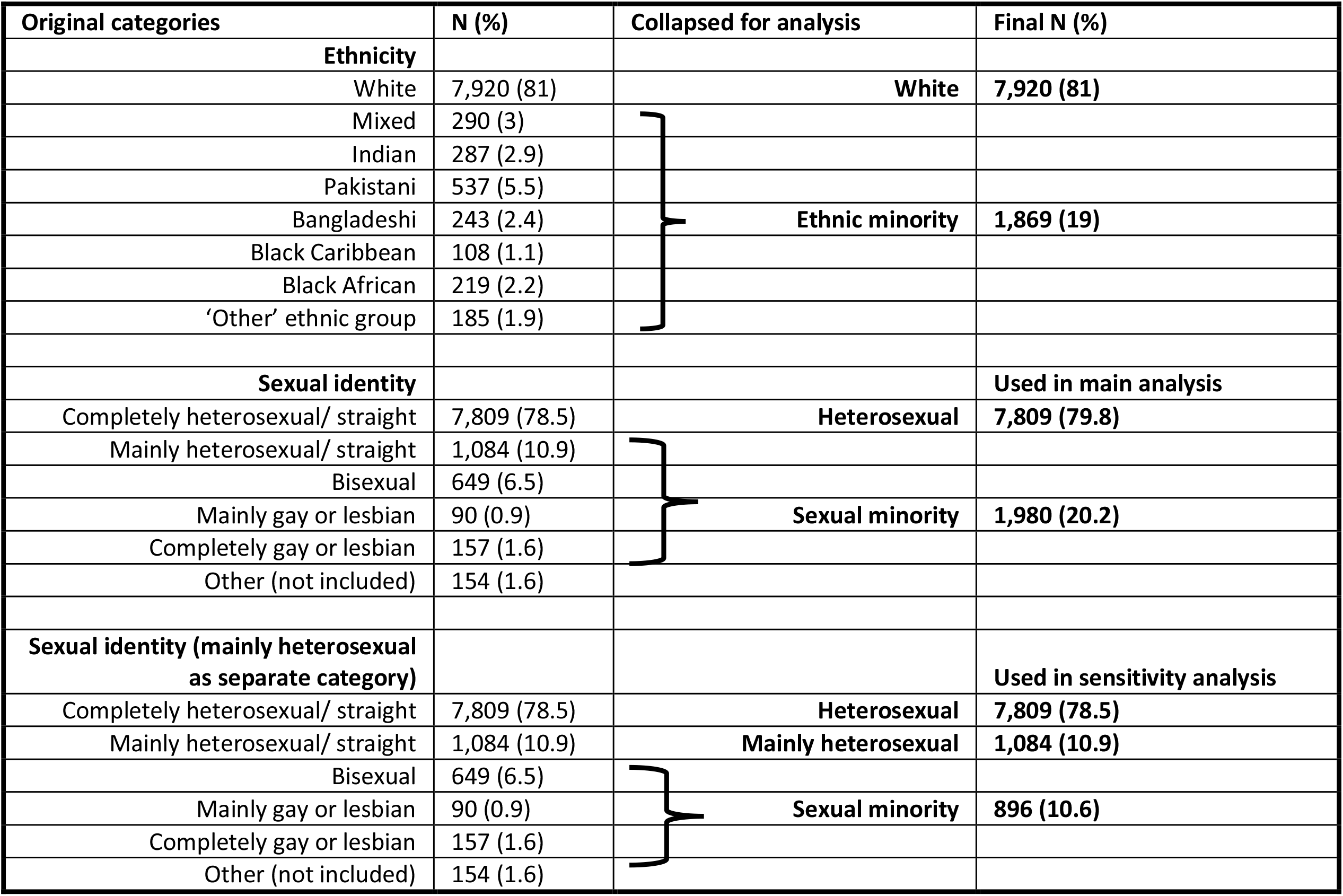
The original categories for ethnicity and sexual identity that were combined for analysis.

Further categorising SM individuals into separate groups (gay, lesbian, and bisexual) was restricted by the small numbers of individuals identifying as ethnic and sexual minority.

As an alternative sexual identity variable, participants were also categorised into 1. Completely heterosexual, 2. Mainly heterosexual and 3. Exclusively sexual minority (bisexual, mainly gay/lesbian and completely gay/lesbian) groups.

Parent/guardian reported participant’s ethnicity at age 3 (original categories in Table 1). Information from subsequent ages was used to replace any missing ethnicity at age 3. For analysis, subjects were grouped into one of two ethnic groups, creating a binary variable: 1.White (ethnic majority) and 2.EM (which included mixed, South Asian, Black and ‘other’). The ‘other’ group included participants from Asia (excluding South Asia), the Middle East and South America. The mixed ethnic group included any combination of mixed-ethnic backgrounds. Supplementary Table S1 displays the distributions of study participants by detailed ethnic and sexual identities.

### Sexuality and ethnicity indicator (study exposure)

The exposure of interest was created by combining the binary sexual and ethnic identity variables resulting in one dual identity variable with the categories: 1. White heterosexual, 2. EM and heterosexual [EM-heterosexual], 3. White and sexual minority [White-SM] and 4. EM and sexual minority [EM-SM]. This approach allows us to estimate risk in all three categories of sexuality- and ethnic-identities compared to the reference White Heterosexual group and incorporates testing for interaction effects between the different categories (and consequently an ‘intercategorical’ approach in examining intersectionality between ethnic and sexual identities and their impact on health).^20^

As a sensitivity analysis with the 3-category coding of sexual identity, we created a second sexuality and ethnicity indicator which included individuals who self-identified as ‘mainly heterosexual’ in a separate category: 1. White heterosexual, 2. EM and heterosexual, 3. White and mainly heterosexual, 4. White and sexual minority [White-SM] and 5. EM and mainly heterosexual and 6. EM and sexual minority [EM-SM]. This was done to examine whether the mainly heterosexual group differed from the totally heterosexual and exclusively sexual minority (lesbians, gays, and bisexual) groups.

### Outcomes of interest Mental and general health

Questionnaires assessed health indicators and health-related behaviors answered by adolescents. Mental health indicators included continuous and binary variables of the Kessler Psychological Distress Scale for nonspecific psychological distress, and the self-report Strengths and Difficulties Questionnaire (SDQ-S) which assesses behavioral markers of mental health difficulties in young people (including its 5 subscales assessing conduct problems, hyperactivity/inattention, emotional problems [depression/anxiety], peer problems and prosocial behavior. The total scores for Kessler and each SDQ-S subscale were categorised into binary variables based on recommended cut-off points to indicate individuals with and without difficulties.

Other mental health indicators included doctor diagnosed depression, self-harm (actions like burning, bruising/pinching, taking an overdose of tablets and pulling out hair), attempted suicide and self-esteem (the 5-item Rosenberg self-esteem scale).

Mental wellbeing was assessed using the shortened Warwick-Edinburgh Mental Wellbeing Scale (WEMWBS) which provides a single summary score indicating overall wellbeing. Internal consistency of the different scales was assessed using Cronbach’s alpha and was found to be acceptable (α>0.7).

General health indicators included self-assessed general health, chronic physical or mental health conditions in the preceding year, quality of sleep and Body Mass Index (BMI, continuous, and categorised into normal vs obesity [including overweight] using the International Obesity Task Force age- and sex-specific cut-offs for 2-18 year-olds).

Social adversity was assessed by experiences of victimisation (i.e., experiences of verbal, physical, sexual assault and/or harassment in the past year).

### Health-related Behaviors

These were coded as binary indicators (never tried/experienced/none vs. yes) and included smoking habits (ever smokers, and current smokers), alcohol consumption, frequency of alcohol consumption in previous 12 months, frequency of binge drinking in previous 4 weeks, any drug use, and specifically cannabis use, and frequency of cannabis use in the previous year. Sexual behavior was assessed by sexual activity (ever had intercourse) and risky sex (did not use any contraception). We also examined frequency of physical activity in the previous week (none vs. any). Antisocial behavior was assessed by one or more of the following acts in the previous 12 months: Pushed or shoved/hit/slapped/punched someone, hit someone with or used a weapon, stolen something from someone, harassed someone via mobile phone/email, sent pictures or spread rumours about someone and made unwelcome sexual approaches/sexually assaulted someone.

The original questions, complete component items of each scale and all health-related indicators, and how they were categorised (including references) are explained in detail in Supplementary Methods and listed in Supplementary Table S3.

### Other Covariates

Parental income was used as an indicator of socioeconomic position. Household income (Organisation for Economic Co-operation and Development UK) was categorised into equalised quintiles (where quintiles 1 and 5 represent the lowest and highest income quintiles respectively). Sex was based on sex at birth.

### Statistical analysis

Associations between the dual sexual-ethnic identity indicator and outcomes of interest were analysed using multivariable linear and logistic regression modelling (for continuous and binary outcomes respectively). To account for the stratified cluster design of the MCS and attrition over time, all regression analyses were weighted with non-response weights from the birth sweep (using the Stata ‘*svy*’ command for survey data). All models were adjusted for sex and parental income. Odds ratios from logistic regression models were plotted for visualisation.

*Sensitivity analysis* – The above regression models were run 1. stratified by sex. 2. with the sexual- and ethnicity-indicator which includes the ‘mainly heterosexual’ group as a separate category.

All analyses were conducted in Stata V16 (StataCorp LP, College Station, Texas).

## Results

The final study sample included 9,789 participants (46.7% female, 20.2% who self-identified as sexual minority, and 9.1% as bisexual, gay or lesbian). 15.9% of the study sample reported high levels of psychological distress based on the K6 psychological distress scale. Relatively high numbers of individuals reported self-harm (22.8%), victimisation (45.2%), and bad quality sleep (31.2%). Sexual minority groups (White and ethnic minority) reported similarly higher proportions of mental health difficulties (32%) based on the K6 psychological distress scale, and more than twice as high as heterosexual peers (Table 2). Based on the SDQ, 40% of White-SM and 32% of EM-SM individuals reported symptoms of psychological distress, substantially higher than White-heterosexual (18.5%) and EM-heterosexual (12.4%) peers. Both sexual minority groups had greater numbers of individuals with hyperactivity and peer problems. Compared to the EM-SM group, the White-SM group were more likely to report doctor diagnosed depression (21.3 vs. 13.1%), self-harm (47.1 vs. 39.9%), mental or physical health conditions (26.3 vs. 20.1%), and attempted suicide (15.6 vs. 12.6%) and victimisation (58.8 vs. 52.2%). White-SM (40.5%) and EM-SM (41.3%) groups reported similarly higher proportions of bad quality sleep and substantially higher than heterosexual peers. EM-SM individuals were more likely to report feeling negative about their weight (63.2 vs. 52%) compared to White-SM individuals.

**Table 2.**
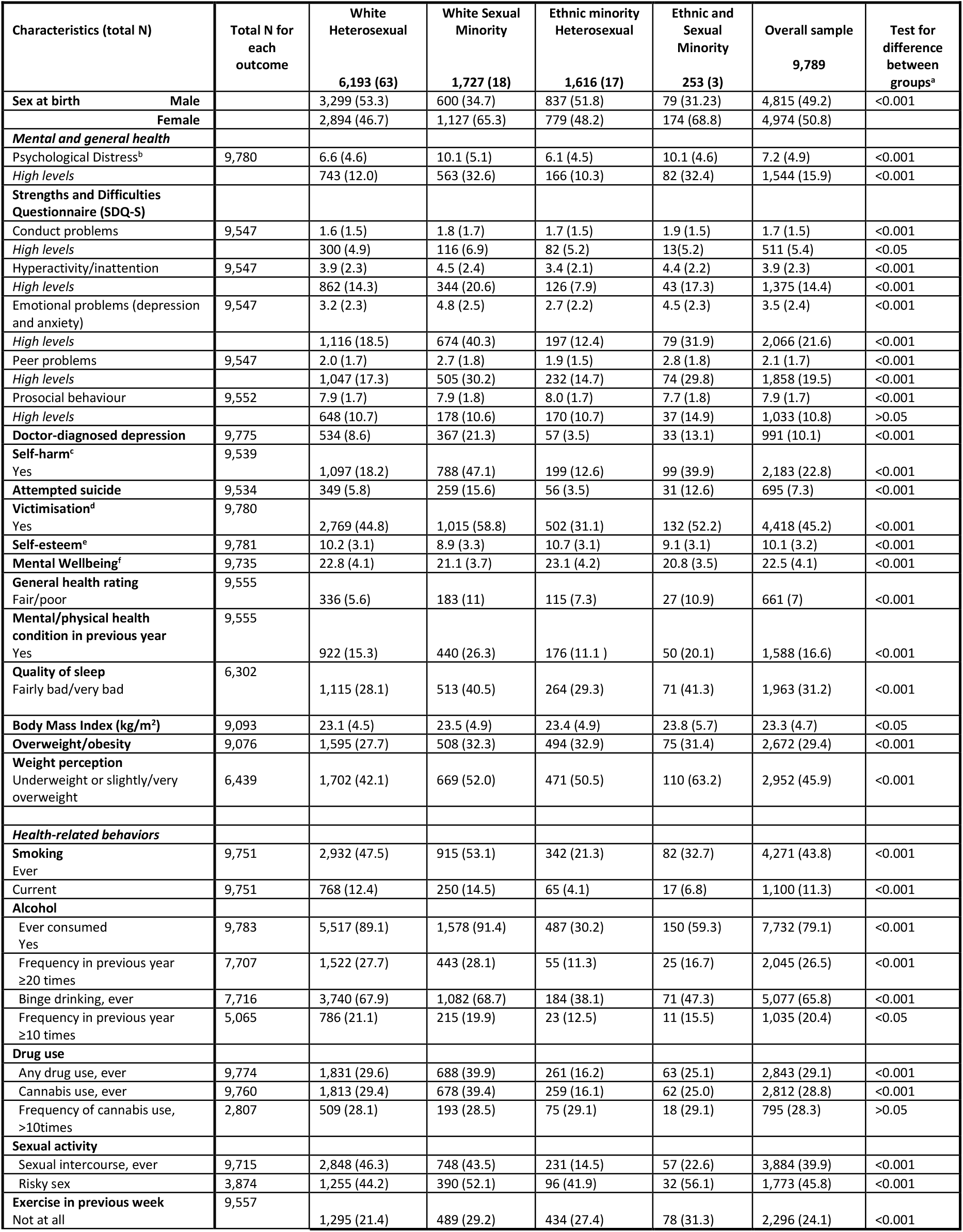
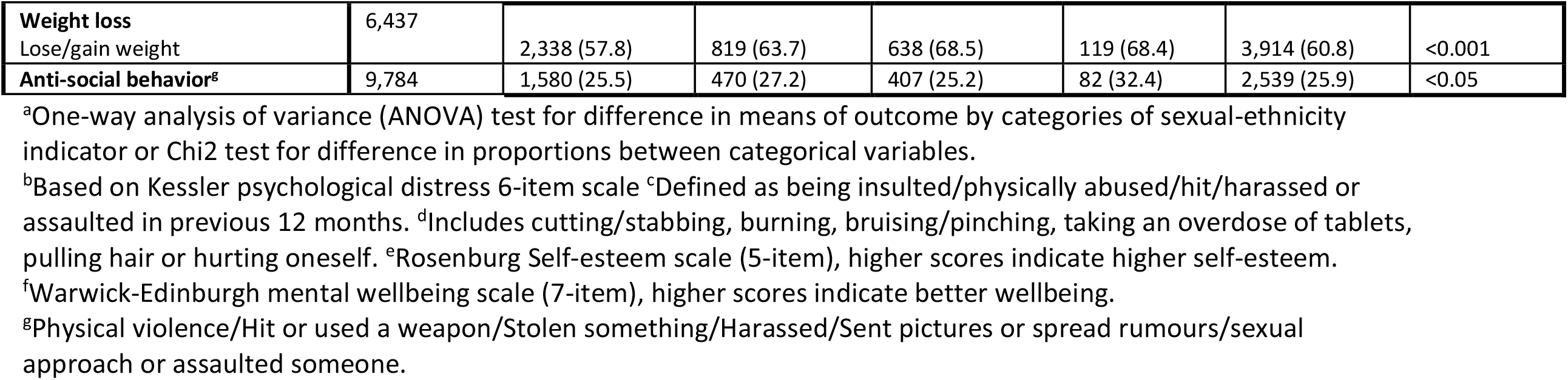
Descriptive characteristics of 9,789 young individuals by ethnic- and sexual-identities that attended the age 17 assessment of the Millennium Cohort Study. Values are means (SD) or N (%)

EM-heterosexual individuals were least likely to report mental health difficulties compared to all other groups (for example, psychological distress: 10.3%, doctor-diagnosed depression: 3.5%, and attempted suicide: 3.5%). Irrespective of sexuality, EM individuals had lower levels of adverse health-related behaviors compared to White individuals (Table 1). However, EM-SM individuals had higher rates of adverse health-related behavior compared to the EM-heterosexual individuals (for e.g., more likely to have tried alcohol (59.3 vs 30.2%), experienced binge drinking (47.3 vs. 38.1%), ever drug use (25.1 vs.16.2%), and risky sex (56.1 vs 41.9%)).

Table 3 and Figure 1 display results from multivariable logistic regression models. Both sexual minority groups had consistently higher odds for mental health difficulties compared to White-heterosexual peers. However, White-SM individuals had higher odds than EM-SM individuals (for example, psychological distress: odds ratio (OR) 3.47[95% CI 2.73-4.42] vs 2.24 [1.35-3.70], self-harm: 3.06[2.41,3.89] vs 1.52[0.98-2.37] and attempted suicide, OR 2.78[2.10-3.68] vs 2.02[1.10-3.71] for White-SM vs EM-SM groups respectively). Only White-SM individuals had higher risks for hyperactivity (OR 1.52[1.22-1.90), doctor diagnosed depression (OR 2.56[1.85-3.53]), poor general health (OR 1.94[1.42-2.64]) and mental/physical health conditions (OR 2.24[1.71-2.94]). Higher odds for prosocial behavior (OR 2.50, 1.35-4.65) was observed in the EM-SM group only.

**Table 3.** Associations between sexual and ethnic identities and binary mental health, wellbeing, and general health outcomes in 9,789 young individuals aged 17 from the Millennium Cohort Study. Estimates are from multivariable logistic regression models (models adjusted for sex and parental income)

|  |  |  | Emotional and behavioral difficulties based on the Strengths and Difficulties Questionnaire |  |  |  |  |  |  |  |  |  |  |  |  |  |
| --- | --- | --- | --- | --- | --- | --- | --- | --- | --- | --- | --- | --- | --- | --- | --- | --- |
| Sexuality and ethnic identity indicator | Psychological distress (K-6)<sup>a</sup> |  | Conduct disorders |  | Hyperactivity |  | Emotional problems |  | Peer problems |  | Prosocial behavior |  | Doctor diagnosed depression |  | Self-harm |  |
|  | OR | 95% CI | OR | 95% CI | OR | 95% CI | OR | 95% CI | OR | 95% CI | OR | 95% CI | OR | 95% CI | OR | 95% CI |
| White-Heterosexual | Ref |  | Ref |  | Ref |  | Ref |  | Ref |  | Ref |  | Ref |  | Ref |  |
| White-SM | 3.47 | 2.73,4.42 | 1.22 | 0.84,1.79 | 1.52 | 1.22,1.90 | 3.17 | 2.48,4.04 | 2.09 | 1.63,2.67 | 1.12 | 0.88,1.41 | 2.56 | 1.85,3.53 | 3.06 | 2.41,3.89 |
| EM-heterosexual | 0.49 | 0.36,0.67 | 0.83 | 0.52,1.31 | 0.51 | 0.37,0.70 | 0.57 | 0.41,0.79 | 0.56 | 0.40,0.78 | 1.06 | 0.73,1.53 | 0.24 | 0.16,0.37 | 0.51 | 0.36,0.72 |
| EM-SM | 2.24 | 1.35,3.70 | 0.72 | 0.39,1.33 | 1.65 | 0.89,3.06 | 1.65 | 1.13,2.40 | 1.38 | 0.85,2.25 | 2.50 | 1.35,4.65 | 1.02 | 0.62,1.67 | 1.52 | 0.98,2.37 |
| N | 9769 |  | 9537 |  | 9537 |  | 9537 |  | 9537 |  | 9542 |  | 9764 |  | 9529 |  |
| Sexuality and ethnic identity indicator | Attempted suicide |  | Victimisation |  | General health |  | Mental/physical health |  | Sleep quality |  | Overweight/obesity |  | Weight perception |  | NA |  |
|  | OR | 95% CI | OR | 95% CI | OR | 95% CI | OR | 95% CI | OR | 95% CI | OR | 95% CI | OR | 95% CI |  |  |
| White-Heterosexual | Ref |  | Ref |  | Ref |  | Ref |  | Ref |  | Ref |  | Ref |  | NA | NA |
| White-SM | 2.78 | 2.10,3.68 | 1.91 | 1.56,2.34 | 1.94 | 1.42,2.64 | 2.24 | 1.71,2.94 | 1.88 | 1.49,2.39 | 1.40 | 1.10,1.78 | 1.65 | 1.32,2.07 |  |  |
| EM-heterosexual | 0.35 | 0.22,0.55 | 0.56 | 0.43,0.75 | 0.91 | 0.62,1.31 | 0.65 | 0.43,0.97 | 1.00 | 0.72,1.40 | 1.07 | 0.86,1.32 | 1.04 | 0.78,1.40 |  |  |
| EM-SM | 2.02 | 1.10,3.71 | 1.30 | 0.89,1.89 | 1.28 | 0.73,2.24 | 1.09 | 0.66,1.79 | 1.67 | 1.01,2.76 | 1.18 | 0.81,1.73 | 1.38 | 0.83,2.29 |  |  |
| N | 9524 |  | 9769 |  | 9545 |  | 9545 |  | 6297 |  | 9066 |  | 6434 |  |  |  |

Text in bold indicate 95% confidence intervals that do not include an odds ratio (OR)=one. EM: Ethnic minority, SM: Sexual minority. <sup>a</sup>Kessler-6 item scale for psychological distress

**Figure 1.**
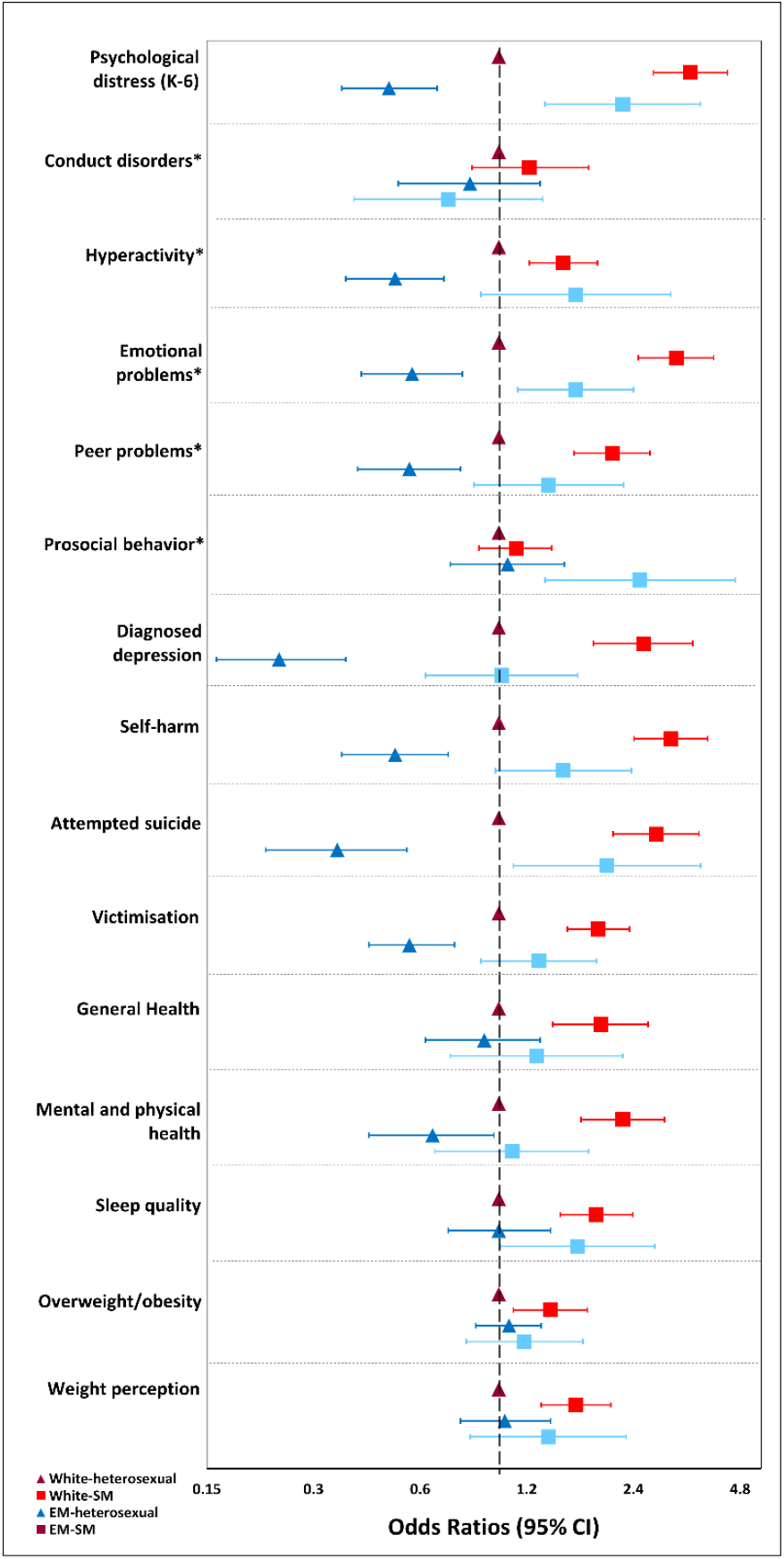
Risk for mental health difficulties, wellbeing and general health based on dual ethnic- and sexual identities in 9,789 individuals aged 17 from the Millennium Cohort Study. Note for Figure 1: *Components of the Strengths and Difficulties Questionnaire (SDQ) CI = confidence interval SM = Sexual minority, EM = ethnic minority

Compared to White-heterosexuals adolescents, White-SM (Table 4, -1.82, [-2.30,-1.33]) and EM-SM (−1.19, [-1.93,-0.44]) groups had worse mental wellbeing.

**Table 4.** Associations between sexuality and ethnic identities and health (continuous outcomes) in 9,789 young individuals aged 17 from the Millennium Cohort Study. Estimates are from multivariable linear regression models (models adjusted for sex and parental income)

|  | Emotional and behavioral difficulties based on the Strengths and Difficulties Questionnaire (SDQ) |  |  |  |  |  |  |  |  |  |  |  |
| --- | --- | --- | --- | --- | --- | --- | --- | --- | --- | --- | --- | --- |
| Sexual and ethnic identity indicator | Psychological Distress (K-6) |  | Conduct problems |  | Hyperactivity |  | Emotional problems |  | Peer problems |  | Prosocial behavior |  |
| | $\beta$ | 95% CI | $\beta$ | 95% CI | $\beta$ | 95% CI | $\beta$ | 95% CI | $\beta$ | 95% CI | $\beta$ | 95% CI |
| White-Heterosexual | Ref |  | Ref |  | Ref |  | Ref |  | Ref |  | Ref |  |
| White-SM | **3.58** | **3.00, 4.15** | **0.15** | **0.02, 0.29** | **0.69** | **0.50, 0.87** | **1.43** | **1.18, 1.68** | **0.69** | **0.48, 0.91** | -0.06 | -0.23, 0.12 |
| EM-heterosexual | **-1.15** | **-1.63, -0.67** | -0.13 | -0.32, 0.06 | **-0.71** | **-0.95, -0.47** | **-0.54** | **-0.77, -0.31** | **-0.39** | **-0.56, -0.22** | 0.18 | -0.01, 0.37 |
| EM-SM | **2.73** | **1.89, 3.56** | **0.29** | **0.06, 0.53** | **0.69** | **0.30, 1.09** | **0.64** | **0.12, 1.17** | **0.48** | **0.15, 0.81** | **-0.49** | **-0.84, -0.14** |
| N | 9769 |  | 9537 |  | 9537 |  | 9537 |  | 9537 |  | 9542 |  |
|  | Self-esteem<sup>a</sup> |  | Mental wellbeing (WEMWBS)<sup>b</sup> |  | Body Mass Index (kg/m<sup>2</sup>) |  | NA |  | NA |  | NA |  |
| | $\beta$ | 95% CI | $\beta$ | 95% CI | $\beta$ | 95% CI | | | | | | |
| White-Heterosexual | Ref |  | Ref |  | Ref |  |  |  |  |  |  |  |
| White-SM | **-1.10** | **-1.40, -0.72** | **-1.82** | **-2.30, -1.33** | 0.36 | -0.22, 0.94 |  |  |  |  |  |  |
| EM-heterosexual | **0.73** | **0.46, 1.00** | **0.90** | **0.43, 1.38** | -0.24 | -0.80, 0.31 |  |  |  |  |  |  |
| EM-SM | **-0.68** | **-1.27, -0.09** | **-1.19** | **-1.93, -0.44** | 0.69 | -0.42, 1.80 |  |  |  |  |  |  |
| N | 9,770 |  | 9724 |  | 9083 |  |  |  |  |  |  |  |

Text in bold indicate 95% confidence intervals that do not include zero. SM: Sexual minority, EM: Ethnic minority, K-6: Kessler-6 item scale for psychological distress,
<sup>a</sup>Rosenberg self-esteem scale (5-item). <sup>b</sup>WEMWBS = Warwick Edinburgh mental wellbeing scale

EM-heterosexual adolescents had consistently and significantly lower odds for all mental health difficulties and general health indicators compared to the White-Heterosexual peers.

Table 5 and Figure 2 display results from multivariable logistic regression models on health-related behaviors. In general, EM individuals were significantly less likely to report adverse health-related behaviors (ever smoking: OR 0.21/0.63, alcohol consumption: OR 0.10/0.25, and ever binge drinking: OR 0.26/0.61 for heterosexual/SM groups respectively) compared to White-heterosexuals individuals. They were also less likely to have ever had sex. In contrast, EM-SM individuals had higher odds for anti-social behavior (OR 1.86 [1.21-2.83]). There were no differences in health-related behaviors between White-heterosexual and White-SM individuals apart from higher odds for drug use (OR 1.45, 1.18-1.78) and risky sex (OR 1.40, 1.03-1.90) in the latter. Compared to the White-heterosexual individuals, the other three groups had higher odds for attempting weight control.

**Table 5.** Associations between sexual and ethnic identities and health-related behaviors in 9,789 young individuals aged 17 from the Millennium Cohort Study. Estimates are from multivariable logistic regression models (models adjusted for sex and parental income)

| Sexual and ethnic identity indicator | Smoking, ever |  | Smoking, regular |  | Alcohol, ever |  | Alcohol frequency |  | Binge drinking, ever |  | Binge drinking, frequency |  | Drug use, ever |  |
| --- | --- | --- | --- | --- | --- | --- | --- | --- | --- | --- | --- | --- | --- | --- |
|  | OR | 95% CI | OR | 95% CI | OR | 95% CI | OR | 95% CI | OR | 95% CI | OR | 95% CI | OR | 95% CI |
| White-Heterosexual | Ref |  | Ref |  | Ref |  | Ref |  | Ref |  | Ref |  | Ref |  |
| White-SM | 1.17 | 0.95,1.45 | 1.10 | 0.80,1.51 | 1.21 | 0.79,1.83 | 1.05 | 0.83,1.33 | 1.18 | 0.96,1.45 | 0.83 | 0.65,1.05 | **1.45** | **1.18,1.78** |
| EM-heterosexual | **0.21** | **0.16,0.27** | **0.18** | **0.12,0.28** | **0.10** | **0.07,0.16** | **0.30** | **0.18,0.51** | **0.26** | **0.18,0.38** | 0.70 | 0.31,1.59 | **0.41** | **0.31,0.55** |
| EM-SM | **0.63** | **0.42,0.94** | 0.69 | 0.31,1.53 | **0.25** | **0.14,0.45** | **0.50** | **0.31,0.83** | 0.61 | 0.37,1.01 | 0.51 | 0.24,1.11 | 1.14 | 0.73,1.77 |
| *N* | 9740 |  | 9740 |  | 9772 |  | 7703 |  | 7712 |  | 5064 |  | 9763 |  |
|  | Cannabis use, ever |  | Cannabis, frequency |  | Sex, ever had |  | Risky sex |  | Exercise |  | Weight control |  | Anti-social behaviour |  |
|  | OR | 95% CI | OR | 95% CI | OR | 95% CI | OR | 95% CI | OR | 95% CI | OR | 95% CI | OR | 95% CI |
| White-Heterosexual | Ref |  | Ref |  | Ref |  | Ref |  | Ref |  | Ref |  | Ref |  |
| White-SM | **1.45** | **1.18,1.78** | 1.18 | 0.86,1.61 | 0.89 | 0.72,1.11 | **1.40** | **1.03,1.90** | 1.22 | 0.97,1.55 | **1.30** | **1.01,1.68** | 1.15 | 0.91,1.44 |
| EM-heterosexual | **0.41** | **0.31,0.55** | 1.40 | 0.81,2.42 | **0.24** | **0.16,0.37** | 0.96 | 0.57,1.60 | 0.96 | 0.73,1.26 | **1.94** | **1.47,2.55** | 0.96 | 0.69,1.33 |
| EM-SM | 1.16 | 0.74,1.81 | **2.65** | **1.05,6.66** | **0.28** | **0.18,0.44** | 1.19 | 0.59,2.41 | 1.4 | 0.94,2.08 | 1.49 | 0.93,2.39 | **1.86** | **1.21,2.83** |
| *N* | 9749 |  | 2805 |  | 9704 |  | 3873 |  | 9547 |  | 6432 |  | 9773 |  |

Text in bold indicate 95% confidence intervals that do not include an odds ratio (OR)=one. EM: Ethnic minority, SM: Sexual minority

**Figure 2.**
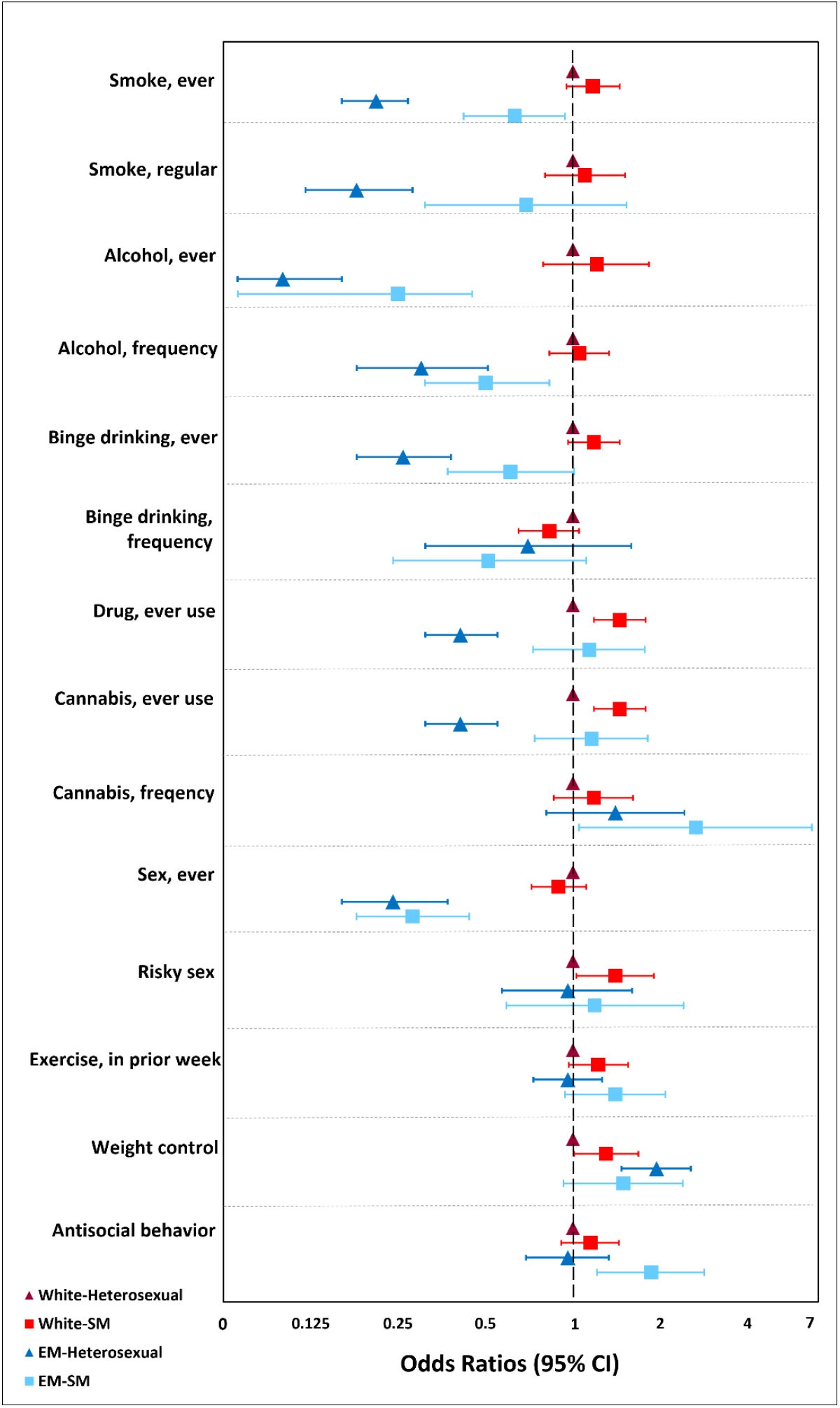
Risk for health-related Behaviors based on dual ethnic- and sexual identities in 9,789 individuals aged 17 from the Millennium Cohort Study. Note for Figure 2: CI = confidence interval SM = Sexual minority, EM = ethnic minority

### Sex stratified results

White SM males and females had increased odds for all outcomes as found in models run with the full sample. The only exceptions were increased odds for conduct disorders (OR 1.91, 1.03-3.54), hyperactivity (OR 1.89, 1.40-2.55), prosocial behaviors (OR 1.62, 1.13-2.33) and worse general health (OR 2.54, 1.72-3.75) in White SM females but not males (Supplementary Table S4). Results were less consistent in EM-SM individuals (for example, significantly higher odds for psychological distress, hyperactivity, prosocial behavior and poor sleep quality in females but not males). Whereas EM-SM males reported significantly higher odds for emotional problems and attempted suicide compared to females (Supplementary Table S4). Similar to models run with the full sample, White-SM individuals were more likely to report adverse health-related behaviors (such as binge drinking and drug use in females) and EM-SM individuals were less likely to report adverse health-related behaviors (Supplementary Table S5). However, White-SM (OR 1.83, 1.23-2.72) and EM-SM (OR 2.33, 1.13-4.81) males were more likely to report weight control. Further, White-SM and EM-SM females were more likely to report cannabis use (ever use and greater frequency) compared to White females.

When examining the mainly heterosexual group separately, the exclusively SM and mainly heterosexual individuals had substantially higher risk for mental health difficulties and poor health, but with a gradient that was more consistent and pronounced in White individuals (for example, OR 2.50 and 4.82 for psychological distress, 2.05 and 5.0 for emotional problems, 1.43 and 4.11 for diagnosed depression, 1.61 and 4.25 for attempted suicide in mainly heterosexual and sexual minority groups respectively, and compared to White-heterosexual individuals, Supplementary Table S6 and Supplementary Figure S1).

This gradient was less evident in EM individuals; the EM-mainly heterosexual group was similar to the EM-Heterosexual group and did not have increased odds for adverse health. However, risk effects for all mental and general health outcomes were substantially larger in effect size in exclusively SM individuals regardless of ethnic origin, when they were analysed as a separate category (Supplementary Table S6). Similarly, the White mainly-heterosexual and EM-mainly heterosexual groups were similar to their heterosexual counterparts in health-related behaviors (Supplementary Table S7).

## Discussion

We report the first population-based estimates for health and health-related behaviors related to dual sexual and ethnic minority identities in a nationally representative sample of adolescents in the UK, revealing: 1. Substantially higher risk for mental health difficulties (consistent with multiple indicators) in SM individuals, regardless of ethnic origin. 2. Adverse health-related behaviors were more commonly reported by White SM individuals and were less likely to be reported by EM individuals (regardless of sexuality). 3. Among White individuals, those who identified as mainly heterosexual had higher odds for mental health difficulties and poor general health that was intermediate between heterosexual and exclusively sexual minority (lesbian, gay and bisexual) groups. This gradient was less evident in mainly heterosexual individuals who are EM. 4. When exclusively SM individuals were examined as a separate group, odds for mental health difficulties, and poor general health were greater, and more so in White individuals.

### Strengths and limitations

Strengths include a large sample size drawn from a nationally representative birth cohort oversampled for EM, and results should be generalizable to the UK population. The study benefits from a wide range of health outcomes, especially multiple and diverse health and wellbeing indicators, and health-related behaviors examined in the same sample. The consistent associations between SM identity and multiple measures of mental health reduces risk of chance findings. Further this study examines risk in EM heterosexual individuals providing an essential comparator group often overlooked in studies.^2^ While all individuals had data on ethnicity, 9% were missing data on sexual identity but proportions missing this information were the same between ethnic groups (9.7% in White and 9.1% in EM). Our approach of using a combined ethnicity and sexuality exposure variable enabled us to test for main effects associated with dual ethnic and sexual identities and interactions between different categories (an intercategorical approach of the intersectionality framework).^20^

Despite a large sample, only 253 (3%) individuals identified as being ethnic and sexual minority (dual minority identities) which precluded analysing differences in health using more specific ethnic (e.g., South Asian, Black and mixed-ethnicity) and SM identities (gay, lesbian and bisexual). This study could not examine SM-specific issues such as age at ‘coming-out’, rejection sensitivity (being rejected by family/close friends), internalised stigma, gender-role strain and perceived burdensomeness which are strongly linked to mental health and wellbeing and are known to differ between ethnic groups.^2, 26^ Similarly, we could not account for immigration status and racial discrimination experiences which may impact health in ethnic minority individuals.

We expected White SM individuals would have significantly higher odds for mental health difficulties and adverse health-related behaviors as per existing evidence. We were unsure of what to expect in EM-SM individuals due to little prior research focussing on this intersection in the UK and European context. Further, EM adolescents in the UK report better mental health compared to their White peers. While we did hypothesise that EM-SM individuals would report worse mental health compared to heterosexual peers, we were unsure if it would be worse than in White-SM peers. To our knowledge, this is the first UK study to examine associations between dual ethnic and sexual identities and diverse health and health-related behaviors in a probability sample. A substantially lower proportion of EM individuals identified as SM (4.6%) and mainly heterosexual (8.9%) compared to White individuals (10.2% and 11.6% respectively). This is comparable with UK national data, and a US study on adolescents, that found EM were more likely to report their sexuality as ‘Other’/’Don’t know’/’refused to answer’ and/or ‘unsure’.^27^ Previous UK studies that examined sexual identity and health focused on mental health and health behaviors excluding ethnic differences.^28, 29^

Most studies that examined associations between ethnic and sexual minority identities and health, have largely focused on adults and are not comparable, as late adolescence is a unique life stage of identity development^2^ These studies largely originate from the US which has a different distribution of EM compared to the UK. Nonetheless, our results are consistent with elevated rates of emotional distress, mood and anxiety disorders, self-harm, and attempted suicide in studies on SM young adults.^2, 30^ In this sample, White-SM individuals had on average higher risk for mental health difficulties, self-harm, and attempted suicide compared to EM-SM individuals but these findings must be interpreted with caution as 95% CIs overlapped, we cannot be certain about differences in risks between the two groups. The lower risk for doctor diagnosed depression in EM groups is consistent with literature suggesting they are less likely to seek clinical support.^31^

Studies on ethnic differences in health-related behaviors among SM adolescents and young adults have mostly focused on risky behaviors like smoking, alcohol, and drug use, with contradictory findings. EM adolescents in the UK, are less likely to smoke, drink, and use drugs compared to their White and mixed-ethnicity peers (though Black adolescents reported higher levels of substance use).^32, 33^ Ethnic and sexual differences in health-related behaviors in UK adolescents are unknown and reported for the first time here – a key contribution of this study. Our findings are intriguing; ethnic- and exclusively SM individuals were less likely to drink but were more likely to use drugs (especially cannabis) compared to their ethnic majority and ethnic minority heterosexual peers. This suggests that EM-SM individuals are to some extent protected by the general lower prevalence of adverse health-related behaviors in EM youth. However, they report some adverse health-related behaviors which are known coping-mechanisms in SM youth. Studies (including a systematic review) indicate that mainly heterosexual individuals experience an intermediate risk for adverse health (between that observed for completely heterosexual and bisexual individuals).^25^ This suggests that mainly heterosexual individuals are a distinct group experiencing worse health outcomes, probably due to experiences of minority stress and non-heterosexual experiences, suggesting that they should be studied as a distinct group where possible^25, 34^

Minority stress theory and the intersectionality framework are two theories put forth to explain the observed worse health and higher rates of health-related risk behaviors in sexual and ethnic minority identified individuals. Minority stress theory is the most cited explanation and considered the foundational framework to explain health disparities in both ethnic and sexual minority individuals.^3^ The theory predicates that minority individuals experience acute and long-term chronic stressors due to social stigma attached to their identities (including racism, heterosexism, victimisation, bullying, unconscious bias, and other forms of discrimination), which are in addition to regular day-to-day stressors experienced by all people. Excess exposure to these forms of minority stress manifest in poorer mental and physical health. This compromised mental health is one of the main predictors of adverse health-related behaviors which further effect health in minority individuals.^35^ Ethnic and SM individuals balance multiple identities (such as gender and/or faith and religious identities) which can further exacerbate stigmatisation and stress associated with having one minority identity.^36^ Additionally, EM individuals encounter a different set of expectations compared to their ethnic majority White peers, including educational, family and community expectations that can be difficult to navigate especially during adolescence and early adulthood. EM-SM individuals face discrimination within LGBTQ+ social contexts adding to feelings of isolation and not being accepted.^37^

Our results suggest it is not the simple accumulation of minority identities that confers increased probability of adverse outcomes, but rather incidents of adversity (or lack of) reflecting a complex interplay of protective and detrimental cultural and interpersonal factors. A substantial body of research has used minority stress theory to explain adverse health and health-related behaviors separately among ethnic and sexual minority groups. The intersectionality theory provides another lens through which we can further understand how unique and multiple social identities (such as ethnicity, gender, sexuality etc.) intersect at the individual-level, reflecting various multiple and reciprocal systems of discrimination that impact health individuals with ≥2minority identities.^3, 19^ Thus, each individual experiences their own unique forms of discrimination and oppression associated with their identities. These social identities are not isolated from one another and simply additive but are interdependent and mutually constitutive.^36^ Thus, rather than conceptualizing stigma-related stress as minority stress experiences which can be broken down as a sum of stress associated with an individual’s identities (e.g., ethnic stress + sexual minority stress), the intersectional framework suggests that different forms of stigma should be tested as multiplicative (i.e., the effects of one form of minority stress are dependent on another form of minority stress).

White-SM and EM-SM individuals experience more frequent and severe victimization, prejudice, and discrimination at individual and institutional levels increasing risk for poor health. But the mechanisms and pathways for adverse health in the two groups potentially occur in different ways, for instance EM-specific and SM specific pathways and unpacking these differences will be important to elucidate mechanisms to reduce health disparities, and potentially design public health interventions accordingly.

The UK’s social and policy contexts are increasingly shifting toward acceptance and equality for sexual minority individuals, yet the current generations of adolescents continue to experience health inequalities.^34^ More effective public health efforts are needed to address continued social stigma and provide access to affirmative care and health promoting resources. Given the complex differences identified at the combination of sexual and ethnic minority identities, public health initiatives are likely to be most effective if differentially designed and targeted with the specific needs of EM communities in mind. Future studies examining health in diverse sexual and ethnic identity groups should analyse protective factors in addition to risk factors. It will be informative to examine how health and health-related behaviors change over time i.e., from adolescence to adulthood and beyond. However, this requires adequately powered longitudinal data including collecting detailed information related to the experiences of sexual and ethnic minority individuals.

## Conclusion

In the UK, SM adolescents (White and EM groups) have substantially higher levels of mental health difficulties (including higher rates of psychological distress, emotional problems, self-harm and attempted suicide) and poorer general health compared to heterosexual peers. Adverse health-related behaviors were more common among White sexual minority individuals compared to EM-SM individuals. Adolescents identifying as mainly-heterosexual had poorer health in between those exclusively heterosexual and exclusively SM. This highlights the need for robust investigation into mechanisms leading to adverse health in sexual minority groups. Additionally, these mechanisms potentially vary between ethnic groups. Further research will benefit from both quantitative and qualitative approaches. Further, the high levels of mental health difficulties experienced by sexual minority individuals highlight the need for accessible and affirming mental health support (for example in schools and universities) with therapists aware of unique risks to health that may be experienced at the intersection of sexual minority and ethnic minority identities.

## Data Availability

All data from the Millennium Cohort Study are anonymised and available to registered researchers via the UK Data Service.

## Author Disclosure Statement section

### Data sharing statement

All data used in this study was collected during the age 17 assessment (MCS 7) of the Millennium Cohort Study. All data is anonymised and is freely available to use for researchers. Data can be downloaded by registering with the UK Data Service. For more information please visit: https://ukdataservice.ac.uk/

For more information on the Millennium Cohort Study including supporting documents such as technical reports and data dictionaries, please see https://cls.ucl.ac.uk/cls-studies/millennium-cohort-study/

## Author Confirmation Statement

All authors confirm that they had full access to all data used in the study and accept responsibility to submit for publication. All authors read and approved the final version of the manuscript. All authors declare no conflicts of interest.

AK conceived the research idea, with input from all authors. AK was responsible for data management and analysis (with input from PP). All authors were involved in interpretation of the findings. AK wrote the original draft (and prepared figures for visualization of findings), and all authors contributed to developing and revising the manuscript. All authors read and approved the final version of the manuscript. AK, VR, DF and PP were involved in funding acquisition for this project

## Acknowledgements

None

## References

1. The health of lesbian, gay, bisexual, and transgender people: building a foundation for better understanding. (IOM (Institute of Medicine), Washington, DC:). 2011.

2. Toomey RB, Huynh VW, Jones SK et al. Sexual minority youth of color: A content analysis and critical review of the literature. J Gay Lesbian Ment Health 2017;21:3–31. DOI: 10.1080/19359705.2016.1217499

3. Meyer IH. Prejudice, social stress, and mental health in lesbian, gay, and bisexual populations: conceptual issues and research evidence. Psychol Bull 2003;129:674–697. DOI: 10.1037/0033-2909.129.5.674

4. Irish M, Solmi F, Mars B et al. Depression and self-harm from adolescence to young adulthood in sexual minorities compared with heterosexuals in the UK: a population-based cohort study. Lancet Child Adolesc Health 2019;3:91–98. DOI: 10.1016/S2352-4642(18)30343-2

5. Mitchell M, Gray G, Green K et al. What works in tackling homophobic, biphobic and transphobic (HBT) bullying among school-age children and young people? (NatCen Social Research). 2014.

6. Russell ST, Sinclair KO, Poteat VP et al. Adolescent health and harassment based on discriminatory bias. Am J Public Health 2012;102:493–495. DOI: 10.2105/AJPH.2011.300430

7. Hatzenbuehler ML, Bellatorre A, Lee Y et al. Structural stigma and all-cause mortality in sexual minority populations. Soc Sci Med 2014;103:33–41. DOI: 10.1016/j.socscimed.2013.06.005

8. Russell ST, Fish JN. Mental health in lesbian, gay, bisexual, and transgender (LGBT) Youth. Annu Rev Clin Psychol 2016;12:465–487. DOI: 10.1146/annurev-clinpsy-021815-093153

9. Saewyc EM. Research on adolescent sexual orientation: development, health disparities, stigma and resilience. J Res Adolesc 2011;21:256–272. DOI: 10.1111/j.1532-7795.2010.00727.x

10. Bhopal RS, Gruer L, Cezard G et al. Mortality, ethnicity, and country of birth on a national scale, 2001-2013: A retrospective cohort (Scottish Health and Ethnicity Linkage Study). PLoS Med 2018;15:e1002515. DOI: 10.1371/journal.pmed.1002515

11. Watkinson RE, Sutton M, Turner AJ. Ethnic inequalities in health-related quality of life among older adults in England: secondary analysis of a national cross-sectional survey. Lancet Public Health 2021;6:e145–e154. DOI: 10.1016/S2468-2667(20)30287-5

12. Khanolkar AR, Amin R, Taylor-Robinson D et al. Inequalities in glycemic control in childhood onset type 2 diabetes in England and Wales-A national population-based longitudinal study. Pediatr Diabetes 2019;20:821–831. DOI: 10.1111/pedi.12897

13. Becares L, Kapadia D, Nazroo J. Neglect of older ethnic minority people in UK research and policy. BMJ 2020;368:m212. DOI: 10.1136/bmj.m212

14. NHS Digital. Mental health of children and young people in England (United Kingdom) 2018.

15. Vitoroulis I, Vaillancourt T. Meta-analytic results of ethnic group differences in peer victimization. Aggress Behav 2015;41:149–170. DOI: 10.1002/ab.21564

16. Galop. The Low Down: Black lesbians, gay men and bisexual people talk about their experiences and needs (United Kingdom) 2001.

17. Bhugra D. Coming out by south Asian gay men in the United Kingdom. Arch Sex Behav 1997;26:547–557. DOI: 10.1023/a:1024512023379

18. Jackson JS, Knight KM, Rafferty JA. Race and unhealthy behaviors: chronic stress, the HPA axis, and physical and mental health disparities over the life course. Am J Public Health 2010;100:933–939. DOI: 10.2105/AJPH.2008.143446

19. Abu-Ras W, Suarez ZE, Breiwish RR. Beyond the axes of inequality: religion, race, and everything in between. Am J Orthopsychiatry 2021;91:217–235. DOI: 10.1037/ort0000478

20. Bauer GR, Churchill SM, Mahendran M et al. Intersectionality in quantitative research: A systematic review of its emergence and applications of theory and methods. SSM Popul Health 2021;14:100798. DOI: 10.1016/j.ssmph.2021.100798

21. Quintana SM, Aboud FE, Chao RK et al. Race, ethnicity, and culture in child development: contemporary research and future directions. Child Dev 2006;77:1129–1141. DOI: 10.1111/j.1467-8624.2006.00951.x

22. Jacob CM, Baird J, Barker M et al. The importance of a life-course approach to health: Chronic disease risk from preconception through adolescence and adulthood - White paper. 2017.

23. Evans-Lacko S, Takizawa R, Brimblecombe N et al. Childhood bullying victimization is associated with use of mental health services over five decades: a longitudinal nationally representative cohort study. Psychol Med 2017;47:127–135. DOI: 10.1017/S0033291716001719

24. Connelly R, Platt L. Cohort profile: UK Millennium Cohort Study (MCS). Int J Epidemiol 2014;43:1719–1725. DOI: 10.1093/ije/dyu001

25. Vrangalova Z, Savin-Williams RC. Psychological and physical health of mostly heterosexuals: a systematic review. J Sex Res 2014;51:410–445. DOI: 10.1080/00224499.2014.883589

26. Pachankis JE, Goldfried MR, Ramrattan ME. Extension of the rejection sensitivity construct to the interpersonal functioning of gay men. J Consult Clin Psychol 2008;76:306–317. DOI: 10.1037/0022-006X.76.2.306

27. Mustanski B, Birkett M, Greene GJ et al. The association between sexual orientation identity and behavior across race/ethnicity, sex, and age in a probability sample of high school students. Am J Public Health 2014;104:237–244. DOI: 10.2105/AJPH.2013.301451

28. Amos R, Manalastas EJ, White R et al. Mental health, social adversity, and health-related outcomes in sexual minority adolescents: a contemporary national cohort study. Lancet Child Adolesc Health 2020;4:36–45. DOI: 10.1016/S2352-4642(19)30339-6

29. Jones A, Robinson E, Oginni O et al. Anxiety disorders, gender nonconformity, bullying and self-esteem in sexual minority adolescents: prospective birth cohort study. J Child Psychol Psychiatry 2017;58:1201–1209. DOI: 10.1111/jcpp.12757

30. Fergusson DM, Horwood LJ, Beautrais AL. Is sexual orientation related to mental health problems and suicidality in young people? Arch Gen Psychiatry 1999;56:876–880. DOI: 10.1001/archpsyc.56.10.876

31. Edbrooke-Childs J, Patalay P. Ethnic differences in referral routes to youth mental health services. J Am Acad Child Adolesc Psychiatry 2019;58:368–375 e361. DOI: 10.1016/j.jaac.2018.07.906

32. Rodham K, Hawton K, Evans E et al. Ethnic and gender differences in drinking, smoking and drug taking among adolescents in England: a self-report school-based survey of 15 and 16 year olds. J Adolesc 2005;28:63–73. DOI: 10.1016/j.adolescence.2004.07.005

33. NHS Digital. Cigarette smoking among 15 year olds - Official Statistics (United Kingdom) 2017.

34. Meyer IH. The elusive promise of LGBT equality. Am J Public Health 2016;106:1356–1358. DOI: 10.2105/AJPH.2016.303221

35. Bontempo DE, D’Augelli AR. Effects of at-school victimization and sexual orientation on lesbian, gay, or bisexual youths’ health risk behavior. J Adolesc Health 2002;30:364–374. DOI: 10.1016/s1054-139x(01)00415-3

36. Khan M, Ilcisin M, Saxton K. Multifactorial discrimination as a fundamental cause of mental health inequities. Int J Equity Health 2017;16:43. DOI: 10.1186/s12939-017-0532-z

37. Ibanez GE, Van Oss Marin B, Flores SA et al. General and gay-related racism experienced by Latino gay men. Cultur Divers Ethnic Minor Psychol 2009;15:215–222. DOI: 10.1037/a0014613

