## Supplemental Methods, Tables and Figure for "Ethnic and sexual identities: inequalities in adolescent health and wellbeing in a national population-based study"

**Supplementary Methods Section – A complete and detailed description of all outcomes of interest**

***Mental and general health***

Information on a range of health indicators was obtained from questionnaires answered by adolescents. The 6 item Kessler Psychological Distress Scale (K6) was used to assess symptoms of nonspecific psychological distress (for example, ''how often do you feel depressed?'') in the preceding 4 weeks[1]. Participants could choose from one of five answers (none of the time, a little of the time, some of the time, most of the time or all the time). Items were scored to create an overall score (higher scores indicate greater levels of depression and anxiety symptoms). The total score was also categorised into a binary variable (with scores ≥13 considered to indicate nonspecific serious psychological distress [SPD] or a high likelihood of having a diagnosable mental illness)[2].

The self-report Strengths and Difficulties Questionnaire (SDQ-S)[3], a brief emotional and behavioural screening tool for young people, consists of 25 items divided into 5 subscales (Supplementary Table 1), was used to assess a range of 'strengths and difficulties' as behavioural markers of mental health difficulties. Each subscale comprises 5 items with a minimum score of 0 (indicating no problems) to a maximum possible score of 10, with higher scores indicating more difficulties. The five subscales assess conduct problems, hyperactivity/inattention, emotional problems (like depression and anxiety), peer problems and prosocial behaviour. Participants reported their behaviour in the previous six months with one of three possible responses: Not true, somewhat true, or certainly true. For each subscale, scores were summed to create an overall score ranging from 0 to 10 (higher scores reflecting higher levels of problems, except for the prosocial subscale, where higher scores indicate fewer difficulties in behaviour). Each of the 5 total scores were then categorised into binary variables based on recommended cut-off points to indicate participants with 'close to average' vs. 'high and very high levels' of difficulties[4].

Other mental health indicators included doctor diagnosed depression, self-harm (self-harm actions included burning, bruising/pinching, taking an overdose of tablets and pulling out hair), attempted suicide and self-esteem (assessed via the 5-item shortened version of the Rosenberg self-esteem scale)[5].

Mental wellbeing was assessed using the shortened 7-item Warwick-Edinburgh Mental Wellbeing Scale (WEMWBS) which provides a single summary score indicating overall wellbeing in preceding 2 weeks[6], and participants could choose one of the following options in response to the seven questions (listed in Supplementary Table 1): None of the time; Rarely; Some of the time; Often; All of the time.

General health was assessed by the question ''How would you describe your health generally?'' with possible answers being excellent, very good, good, fair, or poor. Chronic physical or mental health conditions was assessed by the question ''Do you have any physical or mental health conditions or illnesses lasting or expected to last 12 months or more?''. Quality of sleep was assessed by the question ''During the past month, how would you rate your sleep quality overall?''. Finally, Body Mass Index (BMI) was also used as an indicator of physical health. Height and weight were recorded by trained interviewers using standardized instruments used to calculate BMI (weight/[height2]). BMI values >50kg/m2 and <10kg/m2 were considered implausible and coded as missing. BMI was categorised into normal vs obesity (including overweight) using the International Obesity Task Force age- and sex-specific cut-offs for 2- to 18-year-olds[7].

Social adversity was assessed by experiences of victimisation (i.e., experiences of verbal, physical, sexual assault and/or harassment in the past 12 months).

***Health-related behaviors***

These included smoking habits (ever smokers, and current smokers – yes vs. no), alcohol consumption (never had vs. yes), frequency of alcohol consumption in previous 12 months (<10 times vs. ≥10 times), frequency of binge drinking (≥5 alcoholic drinks at a time) in previous 4 weeks, (<5 times vs. ≥5 time), and any drug use (never vs. yes), and specifically cannabis use (never vs. yes) and frequency of cannabis use in the previous year (≤4 times vs. ≥5times). Sexual behaviour was assessed by sexual activity (ever had intercourse, yes vs. no) and risky sex (i.e., did not use any contraception). We also examined frequency of physical activity in the previous week (none vs. any). Antisocial behaviour was assessed by asking participants if they committed one or more of the following acts in the previous 12 months: Pushed or shoved/hit/slapped/punched someone, hit someone with or used a weapon, stolen something from someone, harassed someone via mobile phone/email, sent pictures or spread rumours about someone and made unwelcome sexual approaches/sexually assaulted someone.

Component items of each scale, all health and wellbeing indicators and how they were categorised are listed in Supplementary Table S2.

**References**

1. Kessler RC, Green JG, Gruber MJ, et al. Screening for serious mental illness in the general population with the K6 screening scale: results from the WHO World Mental Health (WMH) survey initiative. Int J Methods Psychiatr Res 2010; 19 Suppl 1: 4-22.

2. Kim G, DeCoster J, Bryant AN, Ford KL. Measurement Equivalence of the K6 Scale: The Effects of Race/Ethnicity and Language. Assessment 2016; 23(6): 758-68.

3. Goodman R, Meltzer H, Bailey V. The Strengths and Difficulties Questionnaire: a pilot study on the validity of the self-report version. Int Rev Psychiatry 2003; 15(1-2): 173-7.

4. Scoring the Strengths & Difficulties Questionnaire for age 4-17, 2014.

5. Rosenberg M. Society and the adolescent self-image: Princeton University Press; 1965.

6. Tennant R, Hiller L, Fishwick R, et al. The Warwick-Edinburgh Mental Well-being Scale (WEMWBS): development and UK validation. Health Qual Life Outcomes 2007; 5: 63.

7. Cole TJ, Bellizzi MC, Flegal KM, Dietz WH. Establishing a standard definition for child overweight and obesity worldwide: international survey. BMJ 2000; 320(7244): 1240-3.

**Supplementary Table S1. Distribution of 9,789 study participants from the Millennium Cohort Study by the detailed ethnicity and sexual identity. Numbers are N (%)**

| | Sexual identity | | | | |
|---|---|---|---|---|---|
| **Ethnicity** | **Heterosexual** | **Mainly heterosexual** | **Bisexual** | **Gay/Lesbian** | **Total** |
| **White** | 6,193 (78.2) | 917 (11.6) | 584(7.4) | 226 (2.9) | 7,920 |
| **Mixed** | 213 (73.5) | 46 (15.9) | 18 (6.2) | 13 (4.5) | 290 |
| **South Asian** | 968 (90.7) | 64 (6) | 29 (2.7) | 6 (0.6) | 1,067 |
| **Black** | 286 (87.5) | 33 (10.1) | 7 (2.1) | 1 (0.3) | 327 |
| **Other** | 149 (80.5) | 24 (12.9) | 11 (5.9) | 1 (0.5) | 185 |
| **Total** | 7,809 (79.8) | 1,084 (11.1) | 649(6.6) | 247 (2.5) | 9,789 |

**Supplementary Table S2. Distribution of 9,789 study participants from the Millennium Cohort Study by the more detailed ethnic- and sexual-identity predictor variable used in sensitivity analysis. Numbers are N (%)**

| | Sexual identity | | | |
|---|---|---|---|---|
| **Ethnicity** | **Heterosexual** | **Mainly heterosexual** | **Exclusively sexual minority (Lesbian/gay/bisexual)** | **Total** |
| **White** | 6193 (78.2) | 917 (11.6) | 810 (10.2) | 7,920 |
| **Ethnic minority** | 1616 (86.5) | 167 (8.9) | 86 (5) | 1,869 |

**Supplementary Table S3. A detailed description of health and wellbeing outcomes, and health-related behaviours assessed at the age 17 sweep of the Millennium Cohort Study**

| Outcome | Question(s) in cohort member computer-assisted personal interview (CAPI), self-completion interview (CASI) or online questionnaire (CAWI) | Binary or continuous | Comments |
|---|---|---|---|
| **Mental health, wellbeing and general health** | | | |
| General health description | How would you describe your health generally? Would you say it is: | Excellent/very good/good vs fair/poor | |
| Warwick-Edinburg wellbeing scale (short version) | I've been feeling:<br>-optimistic about the future<br>-feeling useful<br>-feeling relaxed<br>-dealing with problems well<br>-thinking clearly<br>-feeling close to other people<br>-able to make up my own mind about things | Continuous | |
| Self-reported Kessler (6 item) | During the last 30 days about how often:<br>- did you feel so depressed that nothing could cheer you up?<br>- did you feel hopeless?<br>- did you feel restless or fidgety?<br>- did you feel that everything was an effort?<br>- did you feel worthless?<br>- did you feel nervous? | Continuous | 1. All of the time<br>2. Most of the time<br>3. Some of the time<br>4. A little of the time<br>5. None of the time |
| Self-reported Strengths and Difficulties Questionnaire (SDQ)<br>-emotional subscale | Complains of headaches/stomach aches/sickness<br>Often seems worried<br>Often unhappy<br>Nervous or clingy in new situations<br>Many fears, easily scared. | Continuous | 1. Not true<br>2. Somewhat true<br>3. Certainly true. |
| -conduct subscale | Often has temper tantrums<br>Generally obedient*<br>Fights with or bullies other children<br>Steals from home, school or elsewhere<br>Often lies or cheats | Continuous | |
| -hyperactivity subscale | Restless, overactive, cannot stay still for long<br>Constantly fidgeting<br>Easily distracted<br>Can stop and think before acting*<br>Sees tasks through to the end*. | Continuous | |
| -peer problems | I am usually on my own. I generally play alone or keep to myself<br>I have one good friend or more *<br>Other people my age generally like me *<br>Other children or young people pick on me or bully me<br>I get on better with adults than with people my own age | Continuous | |
| -prosocial | I try to be nice to other people. I care about their feelings<br>Shares readily with others<br>Helpful if someone is hurt, upset or ill<br>Kind to younger children<br>Often volunteers to help others | Continuous | |
| Rosenberg self-esteem scale (5 item) | How much do you agree or disagree with the following statements about you?<br>On the whole, I am satisfied with myself<br>I feel I have a number of good qualities<br>I am able to do things as well as most other people<br>I am a person of value<br>I feel good about myself | Continuous | |
| Suicidality | Have you ever hurt yourself on purpose in an attempt to end your life? | No vs yes | |
| Doctor diagnosed depression | Has a doctor ever told you that you suffer from depression or serious anxiety? | No vs yes | |
| Mental or physical health condition in previous year | Do you have any physical or mental health conditions or illnesses lasting or expected to last 12 months or more? | No vs yes | |
| Victimisation | In the past 12 months has anyone done any of these things to you?<br><br>Insulted/physical/hit/harassed/assaulted you. | No vs yes (any kind of victimisation) | |
| Self-harm | During the last year, have you hurt yourself on purpose in any of the following ways?<br>Cut or stabbed yourself | No vs yes (any kind of self-harm) | |

| | | | |
|---|---|---|---|
| | Burned yourself<br>Bruised or pinched yourself<br>Taken an overdose of tablets<br>Pulled out your hair<br>Hurt yourself some other way | | |
| Sleep quality | During the past month, how would you rate your sleep quality overall? Would you say it has been…<br>1 …Very good<br>2 …Fairly good<br>3 …Fairly bad, or<br>4 …Very bad? | Very good/fairly good vs fairly bad/very bad | |
| Weight perception (body image) | Which of these do you think you are?<br>1 Underweight<br>2 About the right weight<br>3 Slightly overweight<br>4 Very overweight | Right weight vs overweight/underweight | Which of these do you think you are? |
| Body Mass Index (kg/$m^2$) | | Continuous | |
| Overweight/obesity | | Normal vs. overweight/obesity | |
| Antisocial behaviour | Any antisocial behaviour in previous 12 months:<br>Pushed or shoved/hit/slapped/punched someone?<br>Hit someone with or used a weapon?<br>Stolen something from someone. e.g. a mobile phone, money etc.?<br>Harassed or bothered someone via mobile phone or email?<br>Sent pictures or spread rumours about someone via phone, email, social media or online?<br>Made an unwelcome sexual approach or assaulted someone sexually? | Yes or no | |
| **Health and risky behaviours** | | | |
| Current smoking status | | Never/former versus current | |
| Alcohol use (ever) | Have you ever had an alcoholic drink? That is more than a few sips. | Never vs yes | |
| Frequency of alcohol consumption | How many times have you had an alcoholic drink in the last 12 months? | <5 times vs >5times | |
| Binge drinking | Have you ever had five or more alcoholic drinks at a time? A drink is half a pint of lager, beer or cider, one alcopop, a small glass of wine, or a measure of spirits. | No vs yes | |
| Drug use (any drug use ever) | Have you ever taken any of the following?<br><br>Options included: Cannabis (Marijuana, Dope, Pot, Hash, Grass, Ganja, Weed)<br>Cocaine powder (Coke)<br>Acid or LSD<br>Ecstasy<br>Heroin<br>Crack<br>Speed or Amphetamines<br>Methamphetamine (crystal meth)<br>Ketamine<br>Mephedrone<br>Psychoactive substances | Never vs yes | |
| Cannabis use only (ever) | Have you ever taken any of the following? | Never vs yes | |
| Frequency of cannabis use | In the past year how many times have you taken cannabis? | <4 times vs >4 times | |
| Sexual intercourse (ever had) | Have you ever had sexual intercourse with someone? | No vs yes | |
| Risky sex (use of contraception) | Do you or any partner regularly use any of these forms of contraception or protection when having sex together? | No vs yes | |
| Exercise in previous week | On how many days in the last week did you do a total of at least an hour of moderate to vigorous physical activity?<br>By moderate to vigorous we mean any physical activity that makes you get warmer, breathe harder and makes your heartbeat faster, e.g. riding a bike, running, playing football, swimming, dancing, etc. | None vs. any amount | |
| Weight control via exercise in past year | In the last 12 months, have you exercised to lose weight or to avoid gaining weight | No vs yes | |

**Supplementary Table S4. Associations between ethnic and sexual identities and binary mental and general health outcomes in 9,789 young individuals aged 17 from the Millennium Cohort Study. Estimates are from multivariable logistics regression models run stratified by sex at birth (models adjusted for parental income)**

| | | | Emotional and behavioural difficulties based on the Strengths and Difficulties Questionnaire | | | | | | | | | | | | | |
|---|---|---|---|---|---|---|---|---|---|---|---|---|---|---|---|---|
| ***Males*** | **Psychological distress (K-6)** | | **Conduct disorders** | | **Hyperactivity** | | **Emotional problems** | | **Peer problems** | | **Prosocial behaviour** | | **Doctor diagnosed depression** | | **Self-harm** | |
| | **OR** | **95% CI** | **OR** | **95% CI** | **OR** | **95% CI** | **OR** | **95% CI** | **OR** | **95% CI** | **OR** | **95% CI** | **OR** | **95% CI** | **OR** | **95% CI** |
| **White-Heterosexual** | | | | | | | | | | | | | | | | |
| **White-SM** | **2.99** | **1.89,4.73** | 0.73 | 0.46,1.15 | 1.16 | 0.84,1.60 | **4.88** | **3.15,7.55** | **2.19** | **1.47,3.27** | 0.99 | 0.73,1.35 | **3.11** | **1.63,5.93** | **2.94** | **1.99,4.35** |
| **EM-heterosexual** | **0.47** | **0.28,0.79** | 0.84 | 0.45,1.58 | **0.56** | **0.39,0.81** | 0.65 | 0.36,1.17 | 0.8 | 0.55,1.18 | 0.88 | 0.61,1.27 | **0.34** | **0.17,0.69** | 0.76 | 0.47,1.22 |
| **EM-SM** | 1.78 | 0.83,3.85 | 0.53 | 0.15,1.88 | 0.78 | 0.38,1.59 | **2.43** | **1.23,4.79** | 1.55 | 0.78,3.07 | 1.18 | 0.57,2.45 | 1.29 | 0.47,3.50 | 1.57 | 0.85,2.90 |
| ***N*** | 4805 | | 4716 | | 4716 | | 4716 | | 4716 | | 4720 | | 4804 | | 4720 | |
| ***Females*** | | | | | | | | | | | | | | | | |
| **White-Heterosexual** | | | | | | | | | | | | | | | | |
| **White-SM** | **3.83** | **2.93,5.00** | **1.91** | **1.03,3.54** | **1.89** | **1.40,2.55** | **2.56** | **1.97,3.34** | **2.01** | **1.50,2.70** | **1.62** | **1.13,2.33** | **2.33** | **1.67,3.26** | **3.17** | **2.42,4.16** |
| **EM-heterosexual** | **0.5** | **0.35,0.73** | 0.79 | 0.47,1.32 | **0.46** | **0.27,0.79** | **0.52** | **0.36,0.76** | **0.38** | **0.24,0.59** | 1.74 | 0.85,3.56 | **0.2** | **0.12,0.31** | **0.34** | **0.22,0.51** |
| **EM-SM** | **2.42** | **1.29,4.56** | 1.01 | 0.44,2.32 | **2.2** | **1.02,4.73** | 1.52 | 0.95,2.43 | 1.22 | 0.62,2.39 | **4.4** | **2.17,8.89** | 0.91 | 0.49,1.70 | 1.49 | 0.84,2.63 |
| ***N*** | 4964 | | 4821 | | 4821 | | 4821 | | 4821 | | 4822 | | 4960 | | 4809 | |

| ***Males*** | **Attempted suicide** | | **Victimisation** | | **General health** | | **Mental/ physical health** | | **Sleep quality** | | **Overweight/ obesity** | | **Weight perception** | |
|---|---|---|---|---|---|---|---|---|---|---|---|---|---|---|
| **White-Heterosexual** | | | | | | | | | | | | | | |
| **White-SM** | **4.11** | **2.68,6.28** | **2.31** | **1.68,3.18** | 1.34 | 0.79,2.26 | **1.72** | **1.07,2.77** | **2.28** | **1.53,3.40** | **1.5** | **1.00,2.26** | **2.56** | **1.78,3.68** |
| **EM-heterosexual** | 0.43 | **0.18,1.04** | **0.53** | **0.41,0.69** | 0.96 | 0.53,1.75 | **0.59** | **0.38,0.91** | 0.94 | 0.59,1.49 | 1.1 | 0.82,1.47 | **1.59** | **1.05,2.40** |
| **EM-SM** | **3.6** | **1.46,8.87** | 1.66 | 0.88,3.11 | 1.37 | 0.62,3.05 | 1.16 | 0.59,2.27 | 1.3 | 0.55,3.03 | 0.84 | 0.43,1.67 | 2.13 | 0.85,5.33 |
| ***N*** | 4723 | | 4804 | | 4724 | | 4722 | | 2802 | | 4556 | | 2871 | |
| ***Females*** | | | | | | | | | | | | | | |
| **White-Heterosexual** | | | | | | | | | | | | | | |
| **White-SM** | **2.4** | **1.72,3.36** | **1.72** | **1.33,2.23** | **2.54** | **1.72,3.75** | **2.65** | **1.96,3.58** | **1.65** | **1.23,2.23** | **1.33** | **1.01,1.76** | 1.27 | 0.95,1.68 |
| **EM-heterosexual** | **0.32** | **0.21,0.49** | **0.6** | **0.38,0.93** | 0.84 | 0.58,1.23 | 0.72 | 0.38,1.37 | 1.08 | 0.67,1.72 | 1.03 | 0.70,1.50 | 0.73 | 0.52,1.05 |
| **EM-SM** | 1.68 | 0.89,3.17 | 1.13 | 0.66,1.92 | 1.32 | 0.64,2.72 | 1.15 | 0.62,2.14 | **1.87** | **1.03,3.40** | 1.36 | 0.84,2.21 | 1.06 | 0.59,1.90 |

| | | | | | | | | | | | | | |
|---|---|---|---|---|---|---|---|---|---|---|---|---|---|
| ***N*** | 4801 | | 4965 | | 4821 | | 4823 | | 3495 | | 4510 | | 3563 |

Text in bold indicate 95% confidence intervals that do not include an odds ratio (OR)=one. EM: Ethnic minority, SM: Sexual minority, K-6: Kessler-6 item scale for psychological distress

**Supplementary Table S5. Associations between sexual and ethnic identities and health-related behaviours behaviors in 9,789 young individuals aged 17 from the Millennium Cohort Study. Estimates are from multivariable logistics regression models run stratified by sex at birth (models adjusted for parental income)**

| Sexual and ethnic identity indicator | Smoking, ever | | Smoking, regular | | Alcohol, ever | | Alcohol frequency | | Binge drinking, ever | | Binge drinking, frequency | | Drug use, ever | |
|---|---|---|---|---|---|---|---|---|---|---|---|---|---|---|
| ***Males*** | **OR** | **95% CI** | **OR** | **95% CI** | **OR** | **95% CI** | **OR** | **95% CI** | **OR** | **95% CI** | **OR** | **95% CI** | **OR** | **95% CI** |
| **White-Heterosexual** | 1 | 1.00,1.00 | 1 | 1.00,1.00 | 1 | 1.00,1.00 | 1 | 1.00,1.00 | 1 | 1.00,1.00 | 1 | 1.00,1.00 | 1 | 1.00,1.00 |
| **White-SM** | 1.37 | 0.95,1.96 | 1.23 | 0.65,2.30 | 1.26 | 0.68,2.31 | 1.19 | 0.79,1.81 | 0.85 | 0.59,1.25 | 0.87 | 0.60,1.27 | 1.15 | 0.78,1.67 |
| **EM-heterosexual** | **0.28** | **0.21,0.38** | **0.21** | **0.14,0.32** | **0.14** | **0.09,0.23** | **0.28** | **0.18,0.42** | **0.2** | **0.13,0.29** | **0.54** | **0.31,0.95** | **0.48** | **0.33,0.69** |
| **EM-SM** | **0.45** | **0.25,0.80** | 0.41 | 0.14,1.15 | **0.28** | **0.13,0.62** | 0.63 | 0.29,1.35 | **0.23** | **0.12,0.46** | 1.14 | 0.34,3.78 | **0.46** | **0.24,0.87** |
| ***N*** | 4788 | | 4788 | | 4807 | | 3726 | | 3730 | | 2567 | | 4802 | |
| ***Females*** | | | | | | | | | | | | | | |
| **White-Heterosexual** | 1 | 1.00,1.00 | 1 | 1.00,1.00 | 1 | 1.00,1.00 | 1 | 1.00,1.00 | 1 | 1.00,1.00 | 1 | 1.00,1.00 | 1 | 1.00,1.00 |
| **White-SM** | 1.06 | 0.82,1.37 | 1.01 | 0.72,1.42 | 1.11 | 0.62,1.99 | 0.98 | 0.75,1.27 | **1.46** | **1.14,1.88** | 0.81 | 0.60,1.08 | **1.73** | **1.36,2.20** |
| **EM-heterosexual** | **0.15** | **0.11,0.22** | **0.16** | **0.07,0.35** | **0.07** | **0.04,0.13** | **0.34** | **0.13,0.90** | **0.36** | **0.19,0.68** | 0.91 | 0.22,3.84 | **0.32** | **0.22,0.48** |
| **EM-SM** | 0.67 | 0.42,1.07 | 0.77 | 0.29,2.02 | **0.21** | **0.10,0.45** | **0.42** | **0.21,0.85** | 0.93 | 0.50,1.74 | **0.36** | **0.15,0.88** | 1.68 | 1.03,2.76 |
| ***N*** | 4952 | | 4952 | | 4965 | | 3977 | | 3982 | | 2497 | | 4961 | |

| ***Males*** | Cannabis use, ever | | Cannabis, frequency | | Sex, ever had | | Risky sex | | Exercise | | Weight control | | Anti-social behaviour | |
|---|---|---|---|---|---|---|---|---|---|---|---|---|---|---|
| **White-Heterosexual** | 1 | 1.00,1.00 | 1 | 1.00,1.00 | 1 | 1.00,1.00 | 1 | 1.00,1.00 | 1 | 1.00,1.00 | 1 | 1.00,1.00 | 1 | 1.00,1.00 |
| **White-SM** | 1.1 | 0.75,1.61 | 1.03 | 0.65,1.65 | 0.77 | 0.55,1.08 | 1.01 | 0.68,1.51 | 1.31 | 0.68,2.51 | **1.83** | **1.23,2.72** | 1.44 | 0.95,2.19 |
| **EM-heterosexual** | **0.48** | **0.33,0.70** | 1 | 0.50,1.99 | 0.86 | 0.65,1.14 | **0.39** | **0.27,0.57** | 0.67 | 0.39,1.16 | 0.74 | 0.52,1.04 | **2.21** | **1.48,3.29** |
| **EM-SM** | **0.46** | **0.24,0.88** | 0.83 | 0.25,2.74 | 1.29 | 0.71,2.35 | **0.34** | **0.18,0.63** | 1.94 | 0.72,5.29 | **2.33** | **1.13,4.81** | 0.86 | 0.37,1.99 |
| ***N*** | 4797 | | 1500 | | 4807 | | 4770 | | 1822 | | 4724 | | 2868 | |
| ***Females*** | | | | | | | | | | | | | | |
| **White-Heterosexual** | 1 | 1.00,1.00 | 1 | 1.00,1.00 | 1 | 1.00,1.00 | 1 | 1.00,1.00 | 1 | 1.00,1.00 | 1 | 1.00,1.00 | 1 | 1.00,1.00 |
| **White-SM** | **1.76** | **1.39,2.24** | **1.52** | **1.07,2.15** | **1.82** | **1.39,2.38** | 0.81 | 0.63,1.02 | **1.46** | **1.12,1.91** | 1.02 | 0.77,1.36 | 1.21 | 0.88,1.66 |
| **EM-heterosexual** | **0.33** | **0.22,0.49** | **2.93** | **1.38,6.24** | 1.19 | 0.68,2.10 | **0.14** | **0.07,0.29** | 1.52 | 0.54,4.32 | 1.12 | 0.79,1.58 | **1.69** | **1.20,2.38** |
| **EM-SM** | **1.73** | **1.04,2.87** | **3.77** | **1.30,10.93** | **2.44** | **1.53,3.89** | **0.25** | **0.13,0.46** | 1.06 | 0.46,2.40 | 1.15 | 0.69,1.90 | **1.88** | **1.15,3.07** |
| N | 4952 | | 1305 | | 4966 | | 4934 | | 2051 | | 4823 | | 3564 | |

Text in bold indicate 95% confidence intervals that do not include an odds ratio (OR)=one. EM: Ethnic minority, SM: Sexual minority

**Supplementary Table S6. Associations between ethnic and sexual identities and binary mental and general health outcomes in 9,789 young individuals aged 17 from the Millennium Cohort Study. Estimates are from multivariable logistics regression models using the more detailed ethnic- and sexual-identity predictor (models adjusted for sex and parental income)**

| **Ethnic and sexual identity indicator** | **Psychological distress (K-6)** | | **Hyperactivity** | | **Emotional problems** | | **Peer problems** | | **Doctor diagnosed depression** | |
|---|---|---|---|---|---|---|---|---|---|---|
| | **OR** | **95% CI** | **OR** | **95% CI** | **OR** | **95% CI** | **OR** | **95% CI** | **OR** | **95% CI** |
| **White-Heterosexual** | Ref | | Ref | | Ref | | Ref | | Ref | |
| **White- Mainly Heterosexual** | **2.50** | **1.84,3.39** | 0.94 | 0.71,1.25 | **2.05** | **1.52,2.77** | **1.53** | **1.10,2.13** | **1.43** | **1.05,1.96** |
| **White-SM** | **4.82** | **3.53,6.57** | **2.29** | **1.73,3.01** | **5.0** | **3.63,6.90** | **2.83** | **2.06,3.88** | **4.11** | **2.72,6.21** |
| **EM-heterosexual** | **0.50** | **0.36,0.68** | **0.52** | **0.38,0.70** | **0.57** | **0.41,0.79** | **0.56** | **0.40,0.79** | **0.24** | **0.16,0.37** |
| **EM—Mainly heterosexual** | 1.53 | 0.88,2.67 | 0.77 | 0.47,1.26 | 1.26 | 0.84,1.90 | 1.17 | 0.66,2.09 | 0.59 | 0.28,1.27 |
| **EM-SM** | **4.01** | **2.01,8.00** | **4.07** | **1.74,9.54** | **2.51** | **1.20,5.23** | 1.83 | 0.86,3.92 | 1.89 | 0.97,3.68 |
| ***N*** | 9769 | | 9537 | | 9537 | | 9537 | | 9764 | |

| **Sexual and ethnic identity indicator** | **Self-harm** | | **Attempted suicide** | | **Mental/physical health** | | **Sleep quality** | | **Weight perception** | | **Overweight /obesity** | |
|---|---|---|---|---|---|---|---|---|---|---|---|---|
| | **OR** | **95% CI** | **OR** | **95% CI** | **OR** | **95% CI** | **OR** | **95% CI** | **OR** | **95% CI** | **OR** | **95% CI** |
| **White-Heterosexual** | Ref | | Ref | | Ref | | Ref | | Ref | | Ref | |
| **White- Mainly Heterosexual** | **2.02** | **1.53,2.65** | **1.61** | **1.08,2.41** | **1.56** | **1.09,2.23** | **1.77** | **1.29,2.42** | **1.54** | **1.16,2.05** | 1.28 | 0.92,1.77 |
| **White-SM** | **4.76** | **3.44,6.59** | **4.25** | **3.10,5.84** | **3.19** | **2.24,4.54** | **2.01** | **1.46,2.77** | **1.79** | **1.29,2.49** | **1.55** | **1.14,2.11** |
| **EM-heterosexual** | **0.51** | **0.37,0.72** | **0.35** | **0.22,0.55** | **0.65** | **0.43,0.98** | 1.01 | 0.72,1.40 | 1.05 | 0.78,1.40 | 1.07 | 0.86,1.32 |
| **EM—Mainly heterosexual** | 1.07 | 0.68,1.69 | 1.64 | 0.57,4.73 | 0.68 | 0.37,1.23 | 1.17 | 0.65,2.10 | 1.48 | 0.83,2.63 | 0.99 | 0.61,1.60 |
| **EM-SM** | **2.65** | **1.20,5.84** | **2.71** | **1.31,5.59** | 2.03 | 0.97,4.24 | **3.29** | **1.56,6.92** | 1.22 | 0.46,3.22 | 1.66 | 0.91,3.04 |
| ***N*** | 9529 | | 9524 | | 9769 | | 9524 | | 9764 | | 9066 | |

Text in bold indicate 95% confidence intervals that do not include an odds ratio (OR)=one. SM: Sexual minority, EM: Ethnic minority, K-6: Kessler-6 item scale for psychological distress

**Supplementary Table S7. Associations between ethnic and sexual identities and health related behaviors in 9,789 young individuals aged 17 from the Millennium Cohort Study. Estimates are from multivariable logistics regression models using the more detailed ethnic- and sexual-identity predictor (models adjusted for sex and parental income)**

| **Sexual and ethnic identity indicator** | **Smoking, ever** | | **Smoking, regular** | | **Alcohol, ever** | | **Alcohol frequency** | | **Binge drinking, ever** | | **Drug use, ever** | |
|---|---|---|---|---|---|---|---|---|---|---|---|---|
| | **OR** | **95% CI** | **OR** | **95% CI** | **OR** | **95% CI** | **OR** | **95% CI** | **OR** | **95% CI** | **OR** | **95% CI** |
| **White-Heterosexual** | Ref | | Ref | | Ref | | Ref | | Ref | | Ref | |
| **White- Mainly Heterosexual** | 0.98 | 0.74,1.29 | 0.72 | 0.52,0.98 | 1.12 | 0.64,1.96 | 1.12 | 0.80,1.56 | 1.11 | 0.84,1.46 | 1.26 | 0.99,1.60 |
| **White-SM** | **1.43** | **1.09,1.89** | **1.56** | **1.02,2.38** | 1.31 | 0.75,2.30 | 0.98 | 0.74,1.30 | 1.27 | 0.97,1.68 | **1.70** | **1.27,2.27** |
| **EM-heterosexual** | 0.21 | 0.16,0.27 | 0.18 | 0.12,0.28 | 0.10 | 0.07,0.16 | 0.30 | 0.18,0.51 | 0.26 | 0.18,0.38 | 0.41 | 0.31,0.55 |
| **EM—Mainly heterosexual** | **0.48** | **0.27,0.84** | **0.15** | **0.06,0.41** | **0.24** | **0.13,0.44** | **0.36** | **0.18,0.70** | **0.43** | **0.21,0.88** | 0.70 | 0.37,1.31 |
| **EM-SM** | 1.02 | 0.52,2.00 | 2.1 | 0.83,5.31 | **0.28** | **0.11,0.68** | 0.82 | 0.37,1.80 | 1.14 | 0.54,2.41 | **2.46** | **1.24,4.88** |
| | 9740 | | 9740 | | 9772 | | 7703 | | 7712 | | 9763 | |
| **Sexual and ethnic identity indicator** | **Cannabis use, ever** | | **Sex, ever had** | | **Risky sex** | | **Exercise** | | **Weight control** | | **Antisocial behaviour** | |
| | **OR** | **95% CI** | **OR** | **95% CI** | **OR** | **95% CI** | **OR** | **95% CI** | **OR** | **95% CI** | **OR** | **95% CI** |
| **White-Heterosexual** | Ref | | Ref | | Ref | | Ref | | Ref | | Ref | |
| **White- Mainly Heterosexual** | 1.26 | 0.99,1.59 | 0.87 | 0.66,1.14 | 1.17 | 0.79,1.73 | 0.98 | 0.73,1.31 | **1.43** | **1.07,1.89** | 1.12 | 0.86,1.46 |
| **White-SM** | **1.69** | **1.26,2.26** | 0.92 | 0.68,1.24 | **1.70** | **1.15,2.51** | **1.53** | **1.10,2.13** | 1.18 | 0.81,1.74 | 1.18 | 0.84,1.66 |
| **EM-heterosexual** | **0.41** | **0.31,0.56** | **0.24** | **0.16,0.37** | 0.96 | 0.57,1.61 | 0.96 | 0.73,1.27 | **1.93** | **1.47,2.54** | 0.96 | 0.69,1.33 |
| **EM—Mainly heterosexual** | 0.70 | 0.37,1.34 | **0.14** | **0.09,0.21** | 1.77 | 0.75,4.15 | 1.26 | 0.76,2.09 | 1.16 | 0.64,2.09 | **1.81** | **1.09,2.98** |
| **EM-SM** | **2.55** | **1.28,5.06** | 0.68 | 0.31,1.49 | 0.97 | 0.39,2.42 | 1.68 | 0.84,3.35 | **2.66** | **1.31,5.40** | 1.95 | 0.90,4.23 |
| ***N*** | 9749 | | 9704 | | 3873 | | 9547 | | 6432 | | 9773 | |

Text in bold indicate 95% confidence intervals that do not include an odds ratio (OR)=one, SM: Sexual minority, EM: Ethnic minority

**Supplementary Figure S1. Risk for mental health difficulties, wellbeing and general health based on dual ethnic- and sexual identities in 9,789 individuals aged 17 from the Millennium Cohort Study**

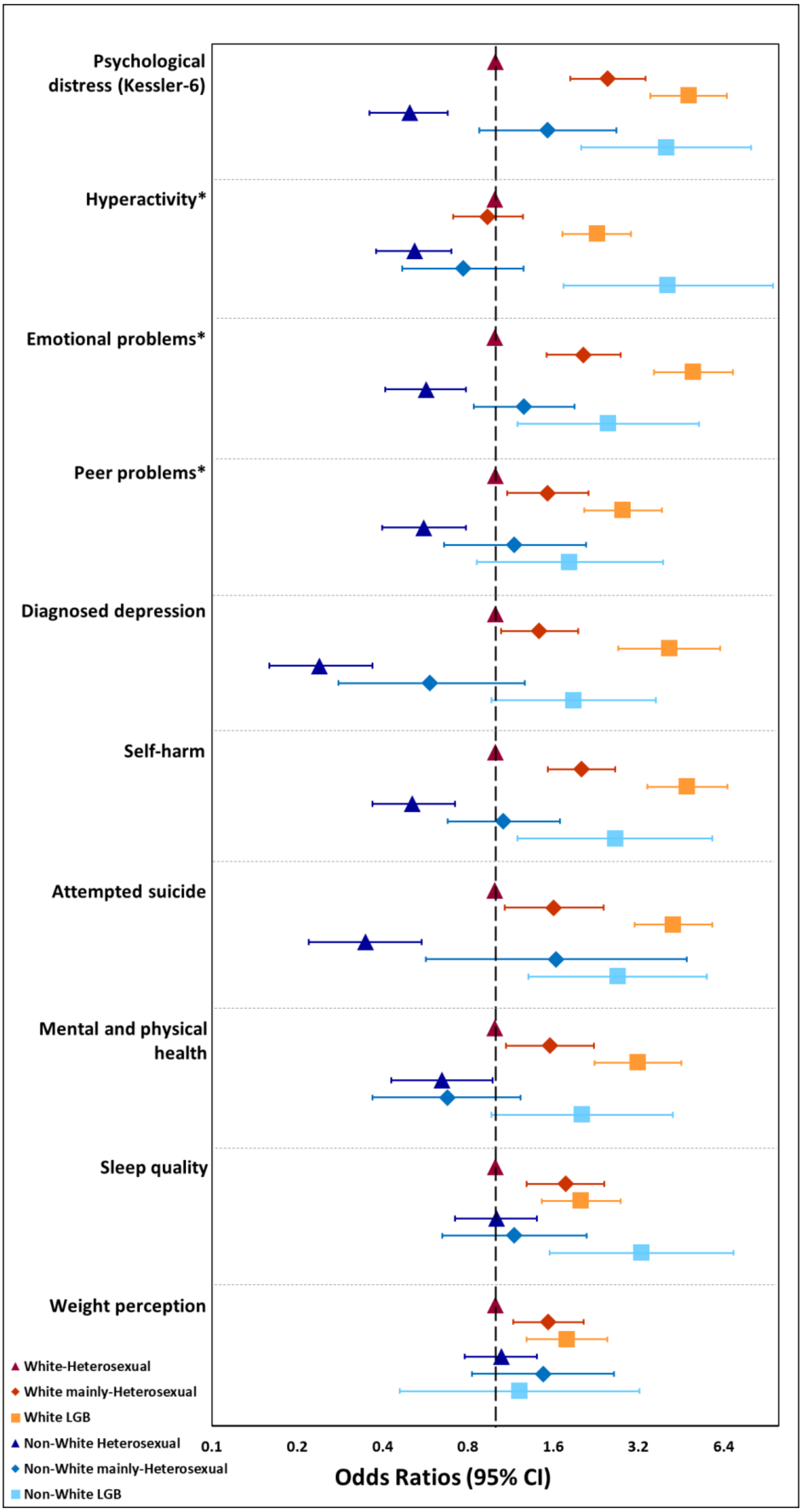

Note for Figure S1:

*Components of the Strengths and Difficulties Questionnaire (SDQ)

CI = confidence interval, LGB = Lesbian, gay and bisexual
